# Modeling the Epidemic Dynamics and Control of COVID-19 Outbreak in China

**DOI:** 10.1101/2020.02.27.20028639

**Authors:** Shilei Zhao, Hua Chen

## Abstract

**Background:** The coronavirus disease 2019 (COVID-19) is rapidly spreading in China and more than 30 countries over last two months. COVID-19 has multiple characteristics distinct from other infectious diseases, including high infectivity during incubation, time delay between real dynamics and daily observed number of confirmed cases, and the intervention effects of implemented quarantine and control measures.

**Method:** We develop a Susceptible, Un-quanrantined infected, Quarantined infected, Confirmed infected (SUQC) model to characterize the dynamics of COVID-19 and explicitly parameterize the intervention effects of control measures, which is more suitable for analysis than other existing epidemic models.

**Results:** The SUQC model is applied to the daily released data of the confirmed infections to analyze the outbreak of COVID-19 in Wuhan, Hubei (excluding Wuhan), China (excluding Hubei) and four first-tier cities of China. We found that, before January 30th, 2020, all these regions except Beijing had a reproductive number *R* > 1, and after January 30th, all regions had a reproductive number *R* < 1, indicating that the quarantine and control measures are effective in preventing the spread of COVID-19. The confirmation rate of Wuhan estimated by our model is 0.0643, substantially lower than that of Hubei excluding Wuhan (0.1914), and that of China excluding Hubei (0.2189), but jumps to 0.3229 after Feb 12th when clinical evidence was adopted in new diagnosis guidelines. The number of un-quarantined infected cases in Wuhan on February 12, 2020 is estimated to be 3,509 and declines to 334 on February 21th, 2020. After fitting the model with data as of February 21th, 2020, we predict that the end time of COVID-19 in Wuhan and Hubei is around late March, around mid March for China excluding Hubei, and before early March 2020 for the four tier-one cities. A total of 80,511 individuals are estimated to be infected in China, among which 49,510 are from Wuhan, 17,679 from Hubei (excluding Wuhan), and the rest 13,322 from other regions of China (excluding Hubei). Note that the estimates are from a deterministic ODE model and should be interpreted with some uncertainty.

**Conclusion:** We suggest that rigorous quarantine and control measures should be kept before early March in Beijing, Shanghai, Guangzhou and Shenzhen, and before late March in Hubei. The model can also be useful to predict the trend of epidemic and provide quantitative guide for other countries in a high risk of outbreak, such as South Korea, Japan, Italy and Iran.

## INTRODUCTION

The outbreak of coronavirus disease 2019 (COVID-19) was initially identified in mid-December 2019 in Wuhan, China [1,2]. The earliest patients in Wuhan are related to exposure from a seafood market. Later, the number of patients grows drastically due to human-to-human transmission [3]. The incubation period of COVID-19 is reported to be 3-7 days, at most 14 days, which varies greatly among patients [2]. The novel coronavirus is believed to be infectious during incubation period when no symptoms are shown on the patients [4], an important characteristics differentiating COVID-19 from its close relative SARS. Considerable measures have been implemented to control the outbreak in Wuhan and China, mainly by quarantine to reduce transmission. On Jan 23rd, 2019, Wuhan restricted travel outside the city. Any person exposed to COVID-19 is required to perform a self-isolation for 14 days. Around Jan 25rd, 2019, nucleic acid kit was developed to diagnose the patients. On Feb 12th, 2020, clinical diagnosis was used to assist the confirmation of infection in Hubei province. Nonetheless, COVID-19 has spread to all provinces of China and more than 30 other countries in the last two months [5,6].

COVID-19 has three features that make it hard to describe with the existing epidemic models including SIR, SEIR etc [7,8]. Firstly, COVID-19 has a relatively long incubation period, which causes a time delay between real dynamic and the daily-observed case numbers. Secondly, the epidemic trend heavily depends on multiple artificial factors, including local medical resources, quarantine measures, and the efficiency of confirmation approaches, which should be explicitly modeled. For example, the outbreak is more severe in Wuhan compared to other cities in China that constrains the medical resources, therefore the infected need a longer time to be confirmed and reported in the official released numbers. This potentially leads to a larger difference between real and reported infected cases in Wuhan than in other places. This could also explain why a sudden increase of confirmed infected cases was observed when clinical diagnosis was adopted in confirmation in Wuhan. Lastly, the quarantine measures are widely implemented, and the quarantined have a lower chance to infect the susceptible individuals. This is critical for controlling the spread across China.

The characteristics of COVID-19 outbreak and control are distinct from existing infectious diseases, and the existing epidemic models cannot be applied to describe the observed data directly. We thus propose to use a simple SUQC model (**S**usceptible, **U**n-quarantined infected, **Q**uarantined infected, **C**onfirmed infected). SUQC distinguishes the infected individuals to be un-quarantined, quarantined but not confirmed, and confirmed. Among the three types, the confirmed number is the data we can directly observe from the official released report. Only un-quarantined infected have ability to infect susceptible individuals and affect the development of the epidemic. In our proposed model, the quarantine rate parameter is used to quantify the strength of quarantine policy on the development of epidemics, and the confirmation rate parameter is used to measure the efficiency of confirmation based on the released data. The two parameters can be solved from fitting the observed confirmed cases over time. Note that the model contains only four variables and three parameters to model both artificial factors and characteristics of epidemics using the data we can directly observe. We expect that the simplified model will not over fit the data given the short time span, but will adequately characterize the essential dynamics.

We apply the SUQC model to the daily released numbers of confirmed cases in Wuhan city, Hubei province (excluding Wuhan), China (excluding Hubei) and four first-tier cities: Beijing, Shanghai, Guangzhou and Shenzhen. The parameters of the model were inferred, and used to predict the future trends of epidemics in China.

## RESULTS

The data of confirmed infected numbers includes 33 consecutive daily records from Jan 20th, 2020 to Feb 21th, 2020 released by the National Health Commission of the People’s Republic of China (see Table S1). The parameters in the model, such as the quarantine rate, are time varying, and thus we divided data into different stages (according to the changes of measures during the epidemic), and we assumed the parameters within each stage are relatively stable. We defined time before Jan 30, 2020 as stage I, and after Jan 30th, 2020 as stage II. To guarantee enough data points within the two stages, the start and end of the stages may vary by one or two days. Wuhan has recently undergone stricter measures of quarantine and transmission limiting, and clinical diagnosis was adopted after Feb 12th, we thus further did a stage III analysis of the dynamics in Wuhan using data after Feb 13th.

### Remarkable difference among trends inferred from three-stage data

Figure 1 shows the inference and prediction of epidemic dynamics of Wuhan using stage I, II and III data respectively. The first 15 daily data points (from Jan 28th to Feb 11th) of stage II were used to fit the model and infer parameters. The following 10 daily data points were used as test data for evaluating the performance of the model. In Figure 1(B), the blue curve presents the model fitting result; blue dots present the predicted numbers of the confirmed infections with the fitted model; and red dots are the observed number of the confirmed infections. Note that the reported numbers of confirmed are based on nucleic acid diagnosis before Feb 12th; both results of nucleic acid and clinical diagnosis are provided from Feb 12th to 14th, and only the total confirmed numbers of the two diagnoses are provided from Feb 15th to 21th. By comparing with three daily data points of the nucleic acid diagnosis, we can see the fitted model predicts the trend well. The predicted numbers of all infected (*I*(*t*)) are also plotted in Figure 1(B). We can see big gap between the predicted numbers of total infections and the predicted numbers of confirmed infections in Wuhan. Clinical diagnosis is adopted by Wuhan local medical agency as an additional diagnosis criterion after Feb 12th, increasing the confirmation rate and causing a big boost of the number of confirmed infections. Note that a proportion of the total infected still remains unidentified even with clinical diagnosis.

**Figure 1.**
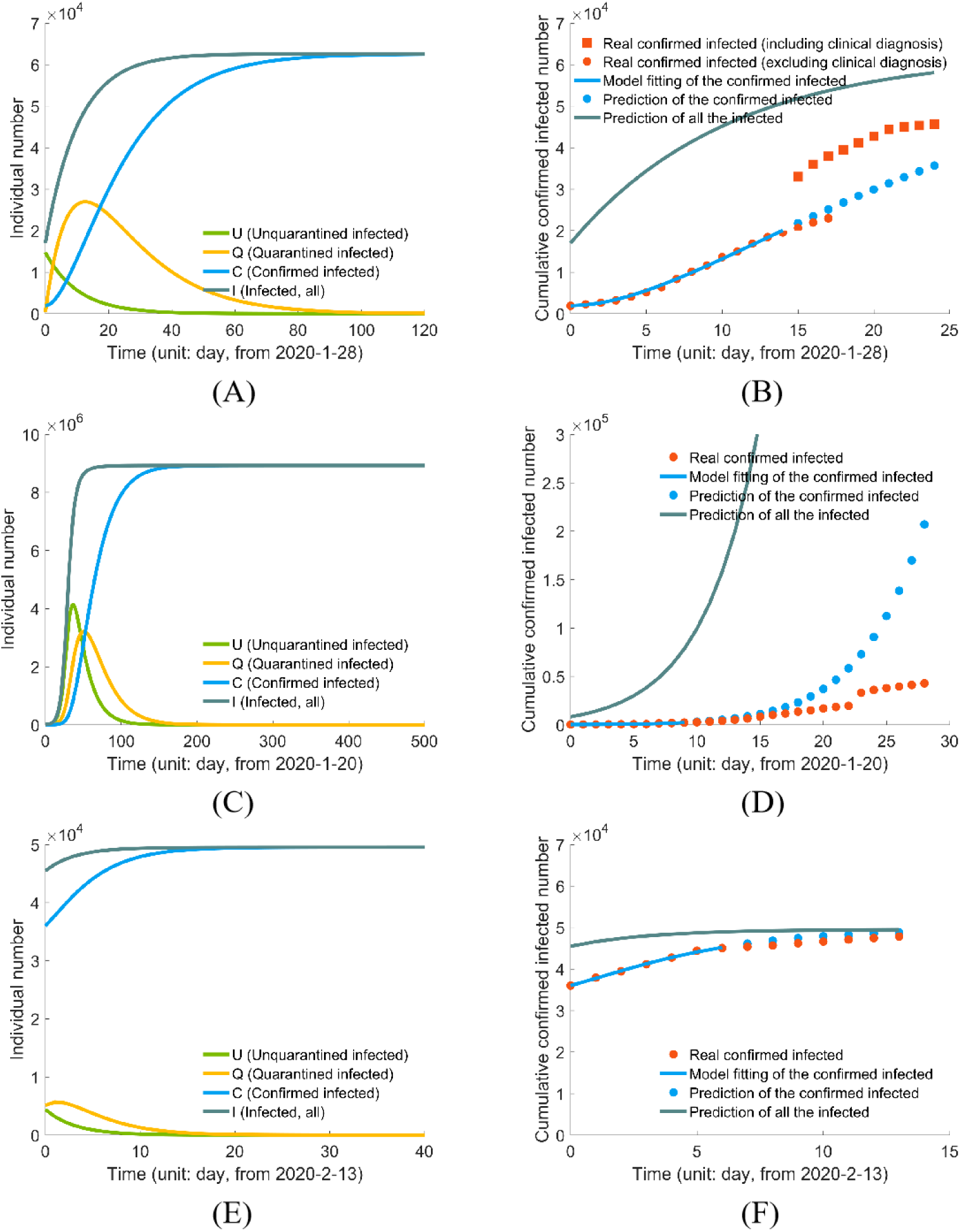
Inferring the epidemic dynamics in Wuhan: (A) prediction using stage II data; (B) Model-fitting and testing with stage II data. The first 15 data points (from Jan 28th) are used to infer the parameters, and the remaining points are used to test the model performance. (C) prediction using stage I data; (D) Model-fitting and testing with stage I data, the first 10 data points (from Jan 20th) are used to infer the parameters, and the remaining points are used to test the model; (E) prediction using stage III data; (F) Model-fitting and testing with stage III data. The first 7 data points (from Jan 23th) are used to infer the parameters, and the remaining points are used to test the model.

With the inferred parameters, we further plot the long-term predictions of the numbers of total infected (*I*), un-quarantined infected (*U*), quarantined infected (*Q*) and cumulative Confirmed infections (*C*) in Wuhan (Figure 1(A)). The end time (increment of confirmed infections equals zero) is predicted to be 147 days from Jan 28th, 2020. The total number of infected individuals is 62,577 (Table 1).

**Table 1.** Parameter estimation of the epidemic dynamics in Wuhan, Hubei (excluding Wuhan) and China (excluding Hubei).

|  | Wuhan |  |  | Hubei (excluding Wuhan) |  | China (excluding Hubei) |  |
| --- | --- | --- | --- | --- | --- | --- | --- |
|  | stageI | stage II | stage III | stageI | stage II | stageI | stage II |
| Quarantine rate | 0.0630 | 0.3917 | 0.6185 | 0.05 | 0.4880 | 0.1941 | 0.5157 |
| Reproductive number | 4.7092 | 0.7575 | 0.4797 | 5.934 | 0.6079 | 1.5283 | 0.5753 |
| Confirmation rate | 0.05 | 0.0643 | 0.3220 | 0.05 | 0.1914 | 0.05 | 0.2189 |
| End time ($U < 1$) | 299 | 101 | 28 | 397 | 47 | 368 | 38 |
| End time ($C_t - C_{t-1} < 1$) | 328 | 147 | 33 | 391 | 55 | 477 | 45 |
| Infected number | 8,923, 823 | 62,557 | 49, 510 | 47,971, 179 | 17,679 | 802,606, 289 | 13,322 |
| $R^2$ | 0.8720 | 0.9746 | 0.9961 | 0.9390 | 0.9863 | 0.9863 | 0.9960 |

**Note the end time is counted from the start point of the stage.**

We can do a similar analysis using stage I data of Wuhan. The stage I data is informative for predicting the epidemic trend assuming no rigorous quarantine and control measures. The first 10 daily data points (from Jan 20th) were used to infer the parameters. The rest data points were used to test the performance of the model (Figure 1(C-D)). As clearly seen in Figure 1(C), the predicted numbers of *C*(*t*) and *I*(*t*) increase dramatically, which are far beyond the observed numbers after Jan 21. The number of total infected can be as large as 8,923,823, and the epidemic lasts for a much longer time (328 days, see Table 1). The dramatic difference between predictions from stage I and stage II data indicates the preventing measures and quarantines, such as travel restrict, are very efficient in controlling the outburst of the epidemic.

Since more strict quarantine and traffic control measures were executed recently to inhibit the infection of COVID-19 in Wuhan and clinical diagnosis was adopted after Feb 12th, we also analyzed the stage III data (from Feb 13th). The estimated quarantine rate is 0.6185, much higher than that of 0.3917 estimated based on stage II data. The total number of infected individuals is estimated to be 49,510, indicating a further acceleration of the epidemic end (Table 1).

Similar analysis was accomplished on stage I and II data of Hubei province (excluding Wuhan), the whole country (excluding Hubei), and four tier-1 cities in China (Figures 2-3, S1-S4, Tables 1-2). Overall, the model predictions are in high accuracy. We see similar trends in these regions: the predicted numbers of infected are distinct between results from the two stage data, indicating the necessity and efficiency of quarantine and control measures. We note that even with stage I data, Beijing have a reproductive number smaller than 1 (Figure S1, Table 2, 0.8840), indicating an early-stage prompt and effective response to COVID-19.

**Table 2.** Parameter inference of the epidemic dynamics in Beijing, Shanghai, Guangzhou and Shenzhen.

|  | Beijing |  | Shanghai |  | Guangzhou |  | Shenzhen |  |
| --- | --- | --- | --- | --- | --- | --- | --- | --- |
|  | stageI | stage II | stageI | stage II | stageI | stage II | stageI | stage II |
| Quarantine rate | 0.3357 | 0.5647 | 0.0817 | 0.5813 | 0.2467 | 0.5838 | 0.05 | 0.5566 |
| Reproductive number | 0.8840 | 0.5254 | 3.6288 | 0.5104 | 1.2026 | 0.5082 | 5.934 | 0.5331 |
| Confirmation rate | 0.0906 | 0.2680 | 0.0881 | 0.2846 | 0.05 | 0.2871 | 0.05 | 0.2599 |
| End time ($U < 1$) | 117 | 19 | 282 | 16 | 481 | 19 | 379 | 17 |
| End time ($C_t - C_{t-1} < 1$) | 103 | 23 | 280 | 20 | 492 | 23 | 372 | 21 |
| Infected number | 814 | 388 | 23,524,072 | 326 | 4,724,899 | 345 | 12,992,106 | 411 |
| $R^2$ | 0.9636 | 0.9839 | 0.9760 | 0.9864 | 0.9290 | 0.9821 | 0.9617 | 0.9845 |

**Note the end time is counted from the start point of the stage.**

**Figure 2.**
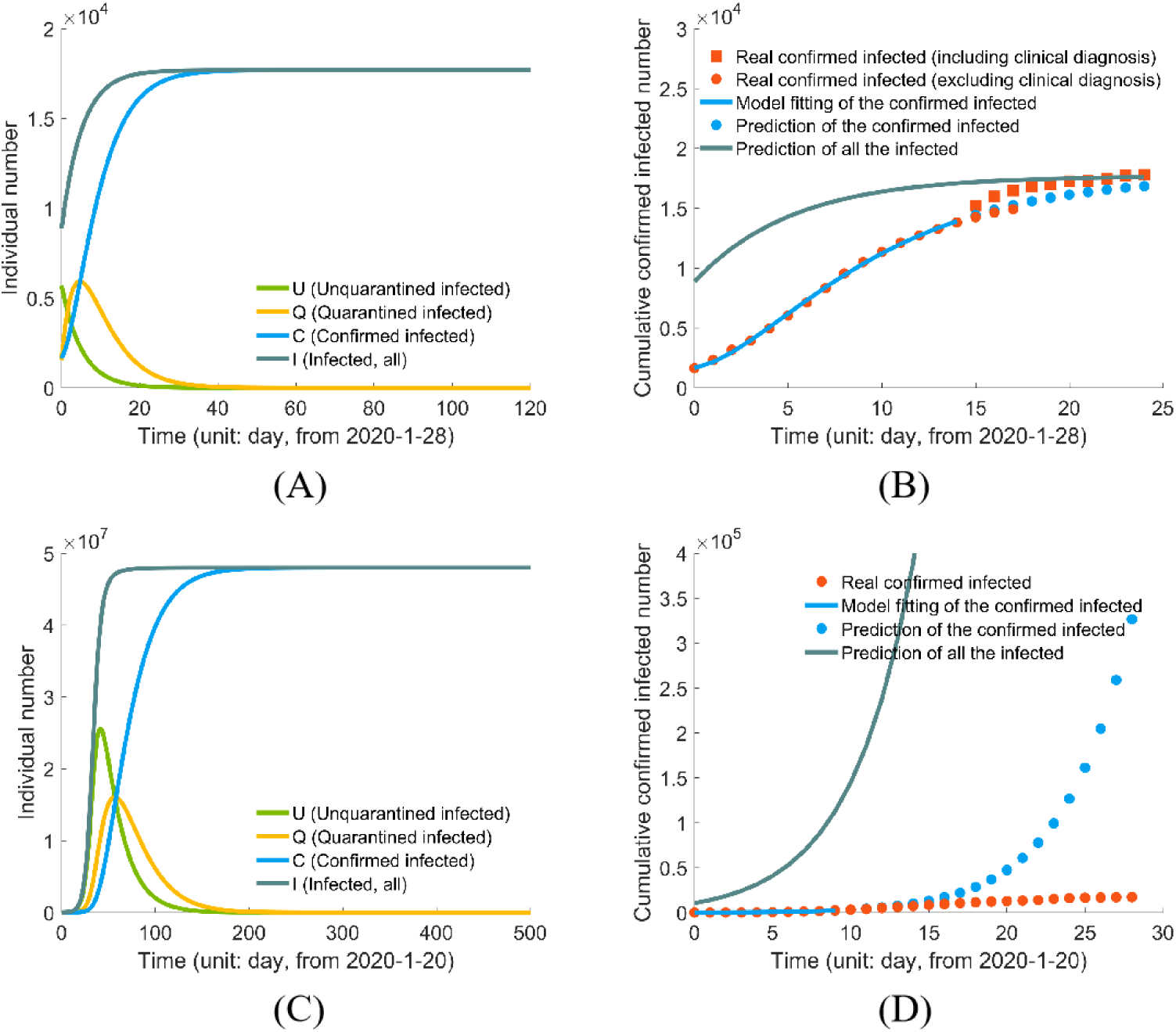
Inferring the epidemic dynamics in Hubei province (excluding Wuhan): (A) prediction using stage II data; (B) model-fitting and testing with stage II data. The first 15 data points (from Jan 28th) are used to infer the parameters, and the remaining points are used to test the model performance; (C) prediction using stage I data; (D) model-fitting and testing with stage I data. The first 10 data points (from Jan 20th) are used to infer the parameters, and the remaining points are used to test the model.

**Figure 3.**
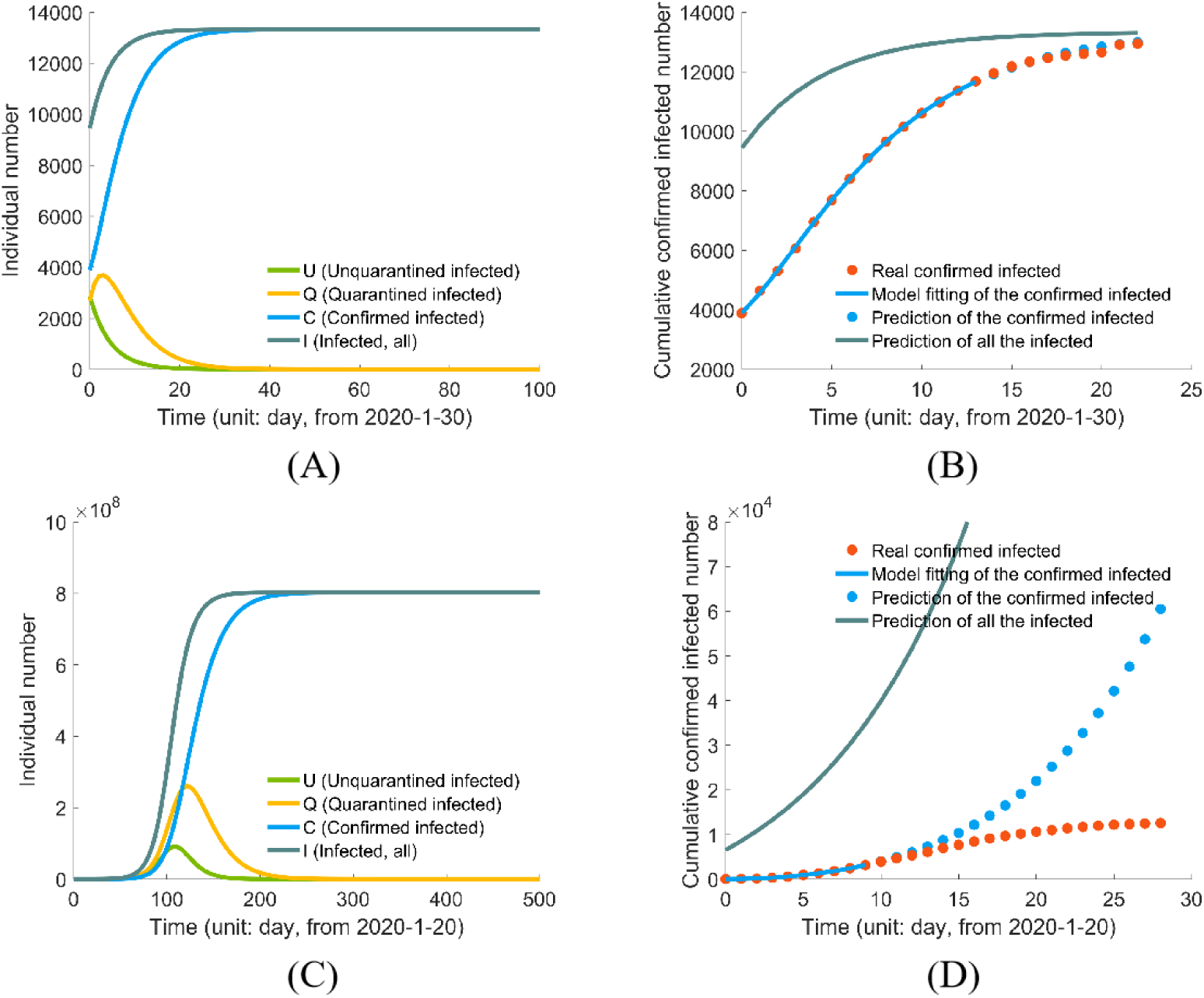
Inferring epidemic dynamics in China (excluding Hubei province): (A) prediction using stage II data; (B) model-fitting and testing with stage II data. The first 15 data points (from Jan 30th) are used to optimize the parameters, and the remaining points are used to test the model; (C) prediction using stage I data; (D) Model-fitting and testing with stage I data. The first 10 data points (from Jan 20th) are used to optimize the parameters, and the remaining points are used to test the model.

### Confirmation rate

From Figures 2-3, S1-S4, we notice that the difference between *C*(*t*) and *I*(*t*) in Wuhan is the biggest. Wuhan has the most infected individuals than any other places in China (more than 50 %), highly beyond the limit of local clinical resources, leading to a long waiting time for confirmation, and the lowest confirmation rate of 0.0643 in Wuhan, compared with 0.1914 of Hubei (excluding Wuhan), 0.2189 of China (excluding Hubei), 0.2680 of Beijing, 0.2846 of Shanghai, 0.2871 of Guangzhou, and 0.2599 of Shenzhen. As predicted by the model, 26,810 quarantined infections haven’t been confirmed in Wuhan by Feb 11th, 2020. The confirmation rate of Wuhan increases to 0.3229 (Table 1) after Feb 12th when clinical diagnosis was adopted in Wuhan.

### Quarantine rate and reproductive number

At stage II, Wuhan has the smallest quarantine rate (0.3917) compared to the other six regions (all close to or larger than 0.5). This leads to a larger reproductive number (*R* = *α* / *γ*_1_). The reproductive number of Wuhan decreases from 0.7575 at stage II to 0.4797 at stage III. Based on reproductive numbers, the epidemic all over China is apparently under control.

### Un-quarantined

The un-quarantined individuals are the source of new infection. Using stage II data, the estimated number of *U*(*t*) individuals in Wuhan (Figure 1(A)) on Feb 12th is still as high as 3,509. The person-to-person transmission will last for more than two months to mid May 2020. However, estimated with stage III data, the quarantine rate of Wuhan increases to 0.6185 and the un-quarantined infected individuals decreases to 334 on Feb 21th, 2020 (Figure 1(E)).

### PREDICTION S

After fitting the model with the recent data from stage II and stage III (for Wuhan), we make a series of predictions about future dynamics of the COVID-19 outbreak in China.

- The end time of the epidemic (with zero new confirmed infections as the criterion) of Wuhan and Hubei (excluding Wuhan) is around late-March, and around mid-March of China. The end time of the four first-tier cities, Beijing, Shanghai, Guangzhou and Shenzhen, is before early March. The end time with zero un-quarantined infections as a criterion is usually earlier than that with zero new confirmed infections.
- The final reported infected number of the whole country is predicted to be 80,511 individuals, among which 49,510 are from Wuhan and 17,679 from Hubei (excluding Wuhan), and the rest 13,322 are from other regions of China.
- Given the inferred end times, rigorous quarantine and control measures should be kept before March in Beijing, Shanghai, Guangzhou and Shenzhen, and before late March in China (including Hubei). We should further point out that, the real confirmed infections in Beijing (Figure S1(B)), Shanghai (Figure S2(B)) and Shenzhen (Figure S4(B)) are a bit larger than the predicted values by our model. This is likely caused by the recent return-to-work tide after the traditional Chinese Spring festival.

## DISCUSSION

We developed a model SUQC for the epidemic dynamics and control of COVID-19. SUQC uses four variables and as few parameters as possible to avoid over-fitting the data, while adequately characterizes the epidemic dynamics. The model is different from the well-known epidemic SEIR model in the following aspects: (1) the infected individuals are classified into un-quarantined, quarantined and confirmed. And only the un-quanrantined can infect the susceptible individuals; While in SEIR, all the infected are infectious; (2) the quarantine rate is a parameter in SUQC to explicitly model the effects of quarantine and control measures; (3) SUQC distinguishes the confirmed infected individuals (observed data) and the total infected individuals, and the parameter confirmation rate is affected by medical resources and the sensitivity of diagnosis methods. Overall, SUQC is developed to characterize the dynamics of COVID-19, and is more suitable for analysis and prediction than adopting existing epidemic models. However, we should emphasize that the estimates of SUQC are from a deterministic ODE model without confidence intervals, and the uncertainty from various sources will be taken into account in future study.

SUQC is applied to the daily released data of China to analyze the dynamics of COVID-19 outbreak, and demonstrates an accurate prediction of the trends with the test data. COVID-19 has currently spread to more than 30 countries. Some countries, such as, South Korea, Japan, Italy and Iran are in their early stage of an outbreak, and the governments are attempting to minimize further spread. SUQC can serve as a useful tool for quantifying parameters and variables concerning the effects of quarantine or confirmation methods on the epidemic, and further provide guidance on the control of the outbreak in these countries.

## METHODS

### SUQC model

SUQC takes into account the following novel epidemic features of COVID-19: (1) the epidemic has an infection probability during the incubation (presymptomatic) period; (2) various isolation measures are used to control the development of the epidemic; (3) the main data source is the daily number of confirmed infections released in the official report, which is affected by the detection method and has a delay between the real infected and confirmed infected number. Four variables related to the features are used to model the flows of people between four possible states:

*S* = *S*(*t*), the number of susceptible individuals with no resistance to disease in the population. *S* is the same as that in existing infectious disease models, e.g. SIR and SEIR.

*U* = *U*(*t*), the number of infected and un-quarantined individuals that can be either presymptomatic or symptomatic. Different from *E* in the SEIR model, *U* are infectious, and can render a susceptible to be un-quarantined infected.

*Q* = *Q*(*t*), the number of quarantined infected individuals. The un-quarantined infected become Quarantined infected by isolation or hospitalization, and lose the ability of infecting the susceptible.

*C* = *C*(*t*), the number of confirmed infected cases. The number of confirmed infections is released by the official agency or media, which may be the only variable with observation that we can access. Note that *C* is usually smaller than the number of real infected individuals, due to the limited sensitivity of diagnosis methods. The duration of incubation can also cause a time delay of confirmation. Nevertheless, *C* is also the number useful for monitoring and predicting the trend of epidemic dynamics.

Besides the aforementioned variables, we have a composite variable *I*(*t*) = *U*(*t*) + *Q*(*t*) + *C*(*t*), representing the real cumulative number of infected individuals at time *t*. The limitation of detection methods and the medical resources can greatly delay the confirmation process, insomuch the confirmation proportion *C* / *I* is less than 1 and time-varying.

*R*, the number of removed individuals, is not included in the model as in the SIR/SEIR models. Once the infected are quarantined, we assume their probability of infecting susceptible individuals is zero, and thus no matter the infected are recovered or not, they have no effect on the dynamics of the epidemic system.

The model comprises the following independent parameters:

*α* is the infection rate, the mean number of new infected caused by an un-quarantined infected per day. *α* ∈[0, ∞).

*γ*_1_ is the quarantine rate for an un-quarantined infected being quarantined, with the range *γ*_1_ ∈[0,1]. The quarantines can be centralized isolation, self isolation, hospitalization and so on. It is a parameter representing multi-resource measures to reduce infection caused by *U*.

*γ*_2_, the confirmation rate of *Q*, is the probability that the quarantined infected are identified to be confirmatory cases by a conventional method, such as the laboratory diagnosis, with the range *γ*_2_ ∈[0,1]. *γ*_2_ is affected by the incubation period duration, medical conditions, accuracy of laboratory tests, and other artificial factors such as the time delay between case confirmation and the official release. *γ*_2_ is time-varying since the change of diagnosis criterion and the improvement of nucleic acid test can accelerate the confirming process.

*σ* is the subsequent confirmation rate of those infected that are not confirmed by the conventional methods, but confirmed with some additional tests. If no other special approaches used, *σ* is set to 0. Combing two sources of confirmation approaches *β* = *γ*_2_ + (1 − *γ*_2_)*σ* is the total confirmation rate.

*δ* is the confirmation rate of the un-quarantined infected who can be identified as confirmed infections without being quarantined.

We thus set up a set of ODE equations to model the dynamics of an infectious disease and the control by artificial factors (Eqn. 1). In the model, *U* goes directly to *C*, or go through *Q* indirectly. Actually, the former can be viewed as a special case of the later with zero delay time during *Q* → *C*. Thus we delete the direct way and simplified the model as Eqn. 2.

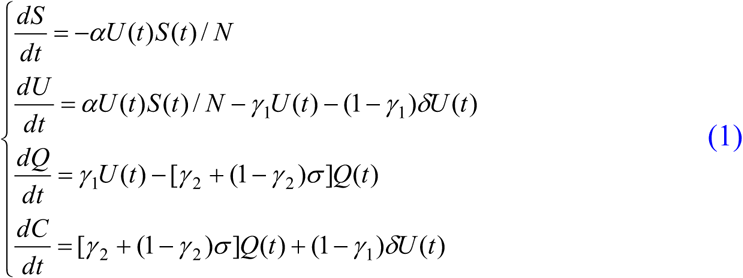

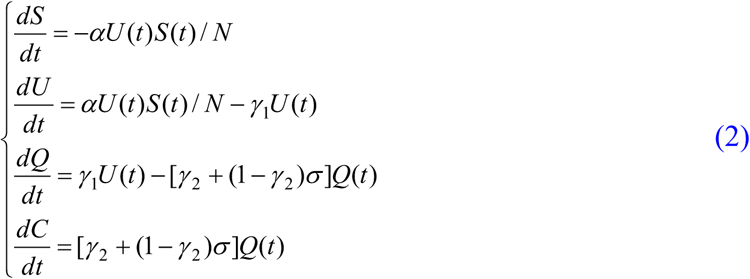

From the above SUQC model, we can further define some biologically meaningful parameters, for monitoring and predicting the trend of disease:

*T* = 1/ *γ*_1_ is the mean waiting time from quarantine to confirmation; *w* = 1/[*γ*_2_ − (1−*γ*_2_)*σ*] is the mean time delay from isolation to confirmation; the reproductive number of the infection is*R* = *α* / *γ*_1_.

### Parameter inference

Among the four variables of the model, the number of cumulative confirmed infections, *C*(*t*), is usually the only variable with daily observed data to be used for model fitting and parameter inference.

The initial value of susceptible individuals *S*(0) is approximately equal to the population size. The initial confirmed infections *C*(0) is the number of infected obtained from the official report. Note that the initial time of the ODE system does not have to be at the beginning of the epidemic, and can start from any time point during the break of COVID-19.

Some parameters can be calculated beforehand using the public data directly. We calculate the infection rate *α* using the confirmed infected numbers of Wuhan city during Jan 20th and Jan 27th. By fitting an exponential curve, we get *α* = 0.2967.

Confirmed infected numbers during this time interval may be less biased and represent natural character of COVID-19, while confirmed numbers are small and fluctuating at early stages before Jan 20th and are affected by strict quarantine measures in later stages. The parameter *α* is hard to estimate accurately. The values estimated by using different methods or different data sets range from 0.3 to 0.5 [9,10,11]. Luckily in SUQC an accurate value of *α* is not necessary; the overall infection ability measured by the reproductive number *R* = *α* / *γ*_1_ as a compound parameter is sensitive in parameter optimization, and thus the bias of *α* can be balanced by *γ*.

Other free parameters and initial values, including *γ*_1_, *β, U*(0) and *Q*(0), are estimated by fitting the daily time series of confirmed infections to the model. Denote *Ĉ* = *f*(*γ*_1_, *β, U*_0_, *Q*_0_) as the expected daily time series of confirmed infections provided by the model (Eqn 2), which was solved by the fourth order Runge-Kutta method with given values of *γ, β, U*(0) and *Q*(0). Define the loss function,

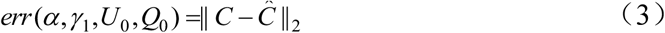

The loss function is then optimized with the interior-point method implemented in the MATLAB function fmincon to infer the parameter values. We try different initial values in parameter optimization, and notice that the inferred parameter values are not sensitive to the provided initial values.

Note that the loss function (Eqn 3) may give too much weight to later observations since the cumulative case numbers are higher than earlier days. We tried another two weighted loss functions to better integrate information across the whole epidemic (Eqns S1 and S2 in the Supplementary information), and compared the prediction of the loss functions. The prediction seems robust on the choice of loss functions (Figures S5 and S6). In practice the loss functions may be chosen by their performances evaluated on the test data.

## Data Availability

the data is from public resources (daily released case numbers by the official agency).
The data is posted in the supplement.

## ACKNOWLEDGEMENTS

We are grateful to Drs. Yongbiao Xue and Liping Wang for motivating this project, to Dr. Hongyu Zhao and the anonymous reviewers for their valuable comments. This project was supported by the National Natural Science Foundation of China (Grant No. 31571370, 91631106, and 91731302), the ‘‘Strategic Priority Research Program’’ of the Chinese Academy of Sciences (Grant No. XDB13000000), the National Key R&D Program of China (Grant No. 2018YFC1406902), and the One Hundred Talents Program of the Chinese Academy of Sciences.

